# Identification of a Novel Alternatively Spliced CRYBA1 Transcript in Unilateral Childhood Cataract Associated with Persistent Fetal Vasculature

**DOI:** 10.64898/2026.07.08.26357271

**Authors:** Sankaranarayanan Rajkumar, Abhay Raghukant Vasavada, Deepa Agrawal, Shail A Vasavada, Vaishali A Vasavada

## Abstract

**Purpose:** To identify transcript-level variants in crystallin genes in paediatric patients with unilateral cataracts.

**Methods:** Anterior capsulorhexis (n=12) from patients underwent surgical management of congenital unilateral cataracts was collected. Total RNA was isolated from lens epithelial cells, and complementary DNA (cDNA) was synthesized. Full-length RNA transcripts of 10 lens-specific crystallin genes were PCR-amplified and analysed via Sanger sequencing. Identified transcript variants were further validated using genomic DNA (gDNA) through Sanger sequencing. In addition, the full-length (∼7,535 bp) CRYBA1 genomic region was sequenced using Oxford Nanopore Technology.

**Results:** Aberrant low molecular weight (LMW) amplicons (∼370 bp) of the CRYBA1 transcript were identified in three patients presented with unilateral cataract. Of 3 patients, 2 had persistent fetal vasculature (PFV) and 1 had pre-existing posterior capsular defect (PPCD). Sanger sequencing revealed a precise loss of exons 2 to 4 in the CRYBA1 RNA transcript. No coding, splice-site, or large deletion variants were detected in the genomic DNA of the patients or their parents. In silico analysis predicted two possible truncated proteins arising from these alternatively spliced transcripts: one comprising the first 11 amino acids of the N-terminal region with a loss of all Greek key motifs, and another comprising 90 amino acids encoded by exons 5 and 6, initiated from an alternative start codon in exon 5, and loss of Greek key motifs 1 & 2.

**Conclusion:** The precise skipping of exons 2 to 4, consistent with canonical splicing signals (5’-GU…AG-3’), in the absence of genomic alterations, suggests the presence of alternatively spliced (AS) CRYBA1 transcripts in human lenses. This is the first report documenting AS-CRYBA1 transcripts in association with childhood cataracts with PFV and PPCD.

## Introduction

Congenital cataract is characterized by varying degrees and patterns of lens opacification in one or both eyes and remains a major global cause of childhood blindness. It affects an estimated 20,000 to 40,000 newborns annually worldwide [1,2]. Although surgical intervention is available, postoperative complications—such as amblyopia, anisometropia, strabismus, impaired stereopsis, and secondary glaucoma—can significantly impact visual development and quality of life, especially in children with unilateral cataract [3].

While bilateral congenital cataracts are frequently associated with known genetic mutations, systemic disorders, or prenatal infections [4,5], the etiology of unilateral cataracts remains largely unclear. Unravelling the molecular mechanisms behind such cases is essential for proper management and prevention.

We believe that the unilateral cataracts might manifest owing to the abnormal molecular mechanisms prevailing in the tissues rather than the germline genetic variants. We hypothesise that germline mutations in lens-expressed genes are more likely to result in bilateral cataracts due to their systemic expression and high penetrance. Conversely, localized, tissue-specific RNA processing events— such as alternative splicing, RNA editing, or transcript-specific mutations—could be associated with unilateral cataracts. It is noteworthy to mention that majority of previous genetic studies have primarily focused on identifying germline variants at the genomic DNA level. However, this approach may have overlooked key post-transcriptional modifications and RNA-level changes, that influence gene expression and protein function in a tissue-specific and developmental context.

Therefore, in this study, we aimed to comprehensively investigate all possible transcript-level variants in the α-, β-, and γ-crystallin gene family. These crystallins constitute over 90% of the total proteins and form the very dense and highly ordered protein matrix of the lens. By combining transcript mutation screening with traditional genomic analysis, we sought to uncover potential molecular alterations at the RNA level that may contribute to the development of unilateral cataract. This approach can reveal pathogenic mechanisms that may not be evident at the genomic level, thereby improving our understanding of the mechanism of unilateral childhood cataract.

### Methodology

This study was approved by the Institutional Ethics Committee (ICIRC/IEC/2012-2015) and was conducted in accordance with the principles of the Declaration of Helsinki. Written informed consent was obtained from the parents or legal guardians of all participants.

### Clinical Evaluation

A comprehensive ophthalmic examination was conducted, including Lea’s visual acuity testing and A-and B-scan ultrasonography. Patients were examined under general anaesthesia, and the type of cataract was assessed intraoperatively using an operating microscope. Only paediatric patients diagnosed with unilateral cataract were included. Exclusion criteria included a family history of congenital cataract (bilateral or unilateral), premature birth, low birth weight, and systemic conditions known to be associated with lens opacification.

### Screening of Crystallin Transcript Variants

Anterior lens capsules containing epithelial cells and aspirated lens material containing epithelial and fiber cells were collected during cataract surgery. Samples were centrifuged at 3000 × g for 10 minutes at 4°C to pellet the cells, which were then washed twice with phosphate-buffered saline (PBS, pH 7.4). Total RNA was extracted using TRIzol reagent according to the manufacturer’s instructions and dissolved in RNase-free water. RNA quantity was measured using a Qubit II Fluorometer with the Qubit RNA BR Assay Kit (Cat. No. Q10210, Life Technologies, CA, USA). First-strand cDNA synthesis was performed using the SuperScript III First-Strand Synthesis System (Cat. No. 18080-51, Life Technologies, CA, USA), following the manufacturer’s protocol. PCR amplification of crystallin gene transcripts was performed in 20 µL reaction volumes containing 1× SapphireAmp Fast PCR Master Mix (Cat. No. RR350, TaKaRa, Shiga, Japan), 50 ng of cDNA, and 0.4–1 µM of forward and reverse primers (Table 1). Thermal cycling conditions are detailed in table 2. Amplicons were verified by agarose gel electrophoresis and subjected to Sanger sequencing. Sequence variants were identified through BLAST analysis.

**Table 1:**
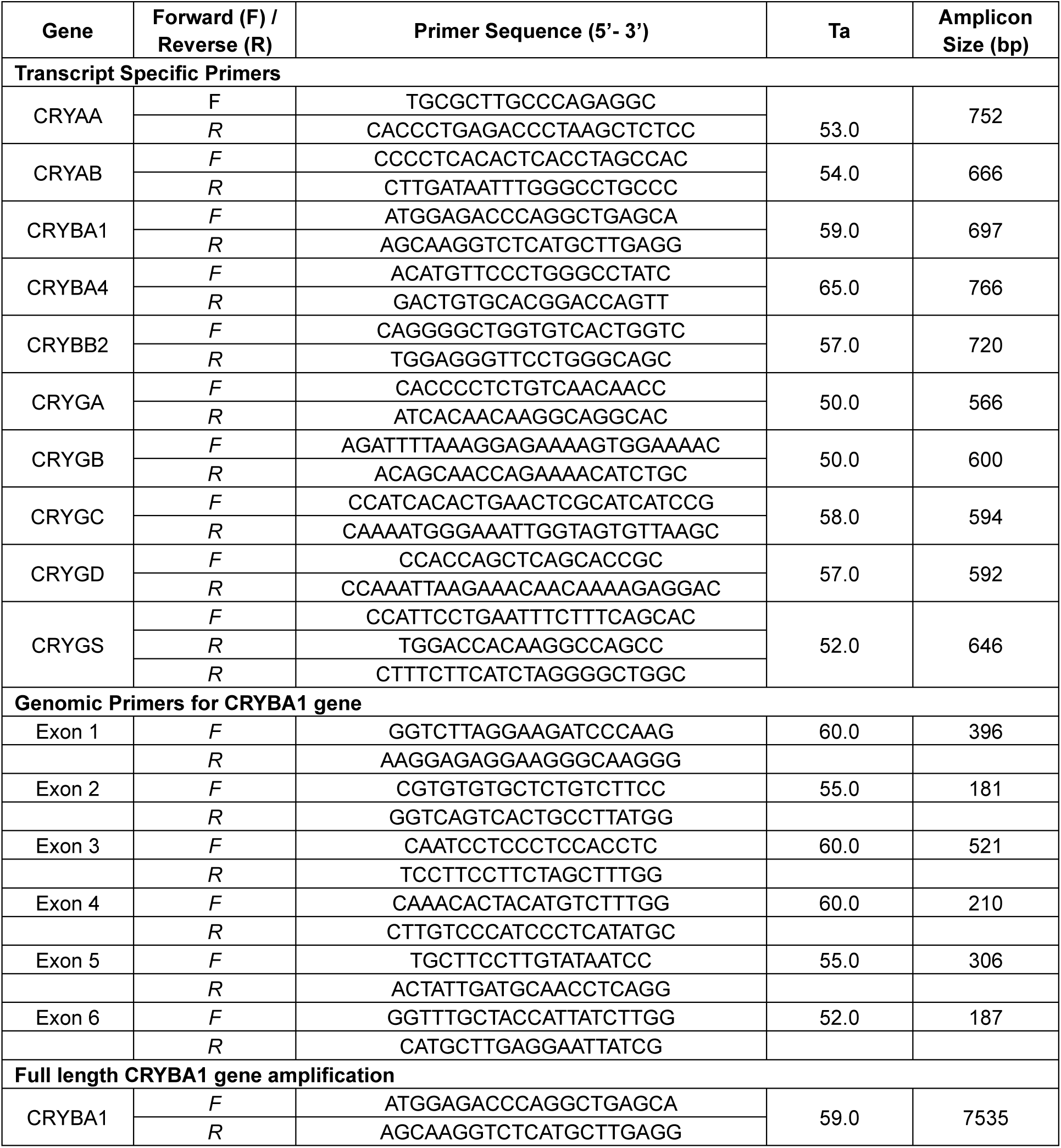
List of primers used for mutation screening.

**Table 2:** PCR Cycling parameters for the amplification of targets.

| **Step** | **Program** | **For Transcript** | **For CRYBA1 Exons** | **For CRYBA1 full gene** |
| --- | --- | --- | --- | --- |
| 1 | Denaturation 1 | 94°C/1min | 98°C/30s | 94°C/3min |
| 2 | Denaturation 2 | 94°C/30sec | 98°C/10s | 94°C/30s |
| 3 | Annealing | 50°C - 62°C/15s | 50°C - 55°C/30s | 57°C/45s |
| 4 | Extension 2 | 72°C/30sec | 72°C/30sec | 65°C/10min |
|  | Cycles (step 2 to 4) | 35 | 28 | 25 |
| 5 | Extension 3 | 72°C/3min | 72°C/2min | 65°C/10min |
| 6 | Cooling | 4°C/∞ | 4°C/∞ | 4°C/∞ |

### Validation of Transcript Variants Using Genomic DNA

Genomic DNA was isolated from peripheral blood samples of patients and available family members using standard protocols [6]. All six exons of the *CRYBA1* gene were amplified using gene-specific primers (Table 1) and sequenced via Sanger sequencing to validate transcript variants.

### Sequencing of CRYBA1 genomic region by Oxford Nanopore Technology (ONT)

To analyse potential large deletions in genomic DNA, the full-length *CRYBA1* gene was amplified using PCR in a 25 µl reaction containing 1X LongAmp PCR Master Mix (Cat. No. M0287L, New England Biolabs, Ipswich, MA, USA), 50 ng of genomic DNA, 10% 360GC enhancer (Cat. No. 4398799, Applied Biosystems, Thermo Fisher Scientific, Waltham, MA, USA), and 2 µM each of forward (5’-ATGGAGACCCAGGCTGAGCA-3’) and reverse (5’-AGCAAGGTCTCATGCTTGAGG-3’) primers. Thermal cycling was performed on a Veriti Thermal Cycler (Applied Biosystems, Thermo Fisher Scientific, Waltham, MA, USA) with the following conditions: initial denaturation at 94°C for 3 minutes; 25 cycles of denaturation at 94°C for 30 seconds, annealing at 57°C for 45 seconds, and extension at 65°C for 10 minutes; followed by a final extension at 65°C for 10 minutes.

The amplified product (∼7535 bp) was confirmed by agarose gel electrophoresis with ethidium bromide staining and purified using the NucleoSpin Gel Extraction Kit (Cat. No. 740609.50, Macherey-Nagel, Düren, Germany).

Approximately 2 µg (25 µl) of purified amplicons were subjected to end-repair and dA-tailing in a 30 µl reaction comprising 3.5 µl of Ultra II End-Prep Reaction Buffer and 1.5 µl of Ultra II End-Prep Enzyme Mix (Cat. No. E7646AA, New England Biolabs, Ipswich, MA, USA). The reaction was incubated at 20°C for 5 minutes, followed by 65°C for 5 minutes. End-prepped products were purified using AMPure XP Beads (Cat. No. A63881, Beckman Coulter, Indianapolis, USA) and eluted in 15 µl of nuclease-free water.

For barcoding, 1 µg (12.5 µl) of each end-prepped amplicon was processed using the Native Barcoding Expansion Kit (Cat. No. EXP-NBD104, Oxford Nanopore Technologies, Oxford, UK) in a 30 µl reaction containing 2.5 µl of Native Barcodes and 15 µl of Blunt/TA Ligase Master Mix (Cat. No. M0367S, New England Biolabs). The mixture was incubated at 25°C for 20 minutes. Equal amounts (250 ng) of barcoded products were pooled, purified with AMPure XP Beads, and eluted in 15 µl of nuclease-free water.

Pooled barcoded amplicons were then ligated with Oxford Nanopore-specific sequencing adapters (AMX) in a 50 µl reaction containing 2.5 µl of AMX II, 5.0 µl of NEB Quick T4 Ligase, 12.5 µl of LNB Buffer (Cat. No. SQK-LSK109, Oxford Nanopore Technologies), and 15 µl of nuclease-free water. The ligation reaction was incubated at 25°C for 10 minutes. The adapter-ligated library was purified using AMPure XP Beads, washed twice with Short Fragment Buffer (SFB, Cat. No. SQK-LSK109) and eluted in 15 µl of Elution Buffer (Cat. No. SQK-LSK109).

For sequencing, 200 fmol (12.0 µl) of the pooled barcoded amplicon library was mixed with 37.5 µl of Sequencing Buffer and 25.5 µl of Loading Beads. The prepared library was stored at −20°C until use.

Flow cell priming was performed with 800 µl of Flush Buffer containing 2.5% Flush Tether through the priming port, with the spot-on port initially closed. An additional 200 µl of Flush Buffer was then loaded into the priming port with the spot-on port open. The sequencing library (75 µl) was subsequently loaded through the spot-on port, and both ports were sealed.

Sequencing was carried out using a MinION device (Oxford Nanopore Technologies) following the standard protocol via MinKNOW software and run for approximately 24 hours at ambient temperature. Raw data were generated in FAST5 format, base-called to FASTQ format, and aligned to the human reference genome (hg38) using Epi2Me. The resulting binary alignment map (BAM) files were sorted, indexed, and visualized using the Integrated Genome Viewer (IGV, version 2.19.2) to identify and validate the genomic variants detected during transcript analysis.

### In silico generation of native and variant CRYBA1 protein models

Protein 3D models for human CRYBA1, native and variants, were built using the full or by deletion of amino acid sequences corresponding to exons 2 to 4 of βA3/A1-crystallin protein, respectively. The construed amino acid sequences were submitted to I-TASSER (Iterative Threading ASSEmbly Refinement) protein structure and function prediction suite for predicting secondary and tertiary structures of the protein [7,8].

## Results

### Demography of the Recruited Subjects

There were 7 males (58.3%) and 5 females (41.7%) with a mean age at which cataract surgery performed was of 1.3 years (range: 0.1 to 4.8 years). Of the 12 patients, 6 (50%) had total, 4 (33.3%) had posterior subcapsular cataracts (PSC), and 1 each had lamellar and sutural cataracts. About 83.3% (n=10) of unilateral cataracts were associated with persistent fetal vasculature (PFV), 66.7% (n=8) were associated with posterior capsular plaque, and 16.7% (n=2) were associated with pre-existing posterior capsular defect. Prenatal TORCH positivity was observed in mothers of 3 probands (Table 3).

**Table 3:** Patient’s demography and ocular complications.

| SID | Gender | TORCH | Affected Eye | Type of Cataract | Presence of PFV | Presence of PCP | Presence of PPCD |
| --- | --- | --- | --- | --- | --- | --- | --- |
| C01 | F | - | OD | PSC | Yes | No | Yes |
| C02 | M | - | OD | Total | No | No | Yes |
| C03 | F | + TXP | OD | Total | Yes | Yes | No |
| C04 | F | + CMV | OD | Total | Yes | Yes | No |
| C05 | F | - | OD | Sutural | No | Yes | No |
| C06 | M | - | OD | Total | Yes | Yes | No |
| C07 | M | - | OS | Lamellar | Yes | No | No |
| C08 | F | +RB | OS | Total | Yes | No | No |
| C09 | M | - | OS | PSC | Yes | Yes | No |
| C10 | M | - | OD | Total | Yes | Yes | No |
| C11 | M | - | OS | PSC | Yes | Yes | No |
| C12 | M | - | OS | PSC | Yes | Yes | No |

F: female, M: male; Sx: Surgery; TORCH: serological test for detecting immunoglobulins (IgG) against Toxoplasmosis-Rubella-Cytomegalovirus and Herpes Simplex Virus. +: test positive, -: test negative, CMV: Cytomegalovirus, RB: Rubella, TXP: Toxoplasmosis); OD: Right Eye, OS: Left Eye; PSC: posterior subscapular cataract, PFV: Persistent Fetal Vasculature; PCP: posterior capsular plaque; PPCD: Pre-existing Posterior Capsular Defect.

### Possibility of Alternatively Spliced CRYBA1 Transcripts in Lens

We observed an aberrant low molecular weight (LMW) amplicon with a size of ∼370 bp (yellow arrows), compared to the control CRYBA1 RNA transcripts (∼697 bp) (Fig. 1A), in 3 patients (C02, C09, C10) (Fig. 1B). Sequencing and subsequent Basic Local Alignment Search Tool (BLAST) analysis of the LMW amplicon (∼370 bp) showed adjoining of exon 1 and exon 5 nucleotide sequences (exon1-exon5 junction), demonstrating a loss of exons 2 to 4 (Fig. 1C), while sequencing of the control amplicon (697 bp) showed continuity of exons from 1 through 6 (Fig. 1D).

**Figure 1:**
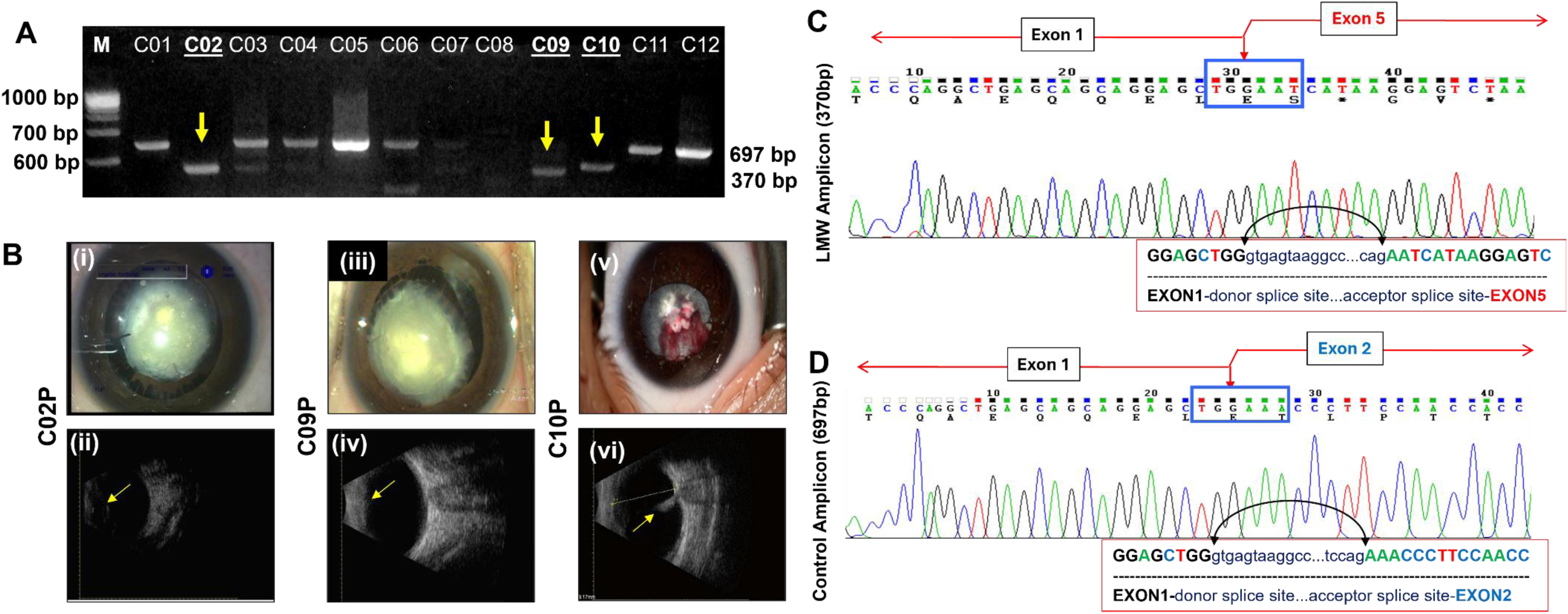
A) Agarose gel electrophoresis of CRYBA1 transcript amplicons shows aberrant low molecular weight (LMW) products (∼370 bp, indicated by yellow arrows) in three patients, compared to the control amplicon (697 bp); M: 100 bp DNA Molecular Weight Marker B) Clinical phenotypes of probands (C02P, C09P, C10P) harbouring LMW CRYBA1 transcripts are presented. (i), (iii), (v) Intra operative assessment of phenotypes & (ii), (iv), (vi) B-scan ultrasonography of posterior segment showing PPCD (ii) and PFV (iv, vi). C02P presented with a unilateral total cataract in the right eye (i) along with a pre-existing posterior capsular defect (PPCD) (ii). C09P exhibited posterior subcapsular cataract in the right eye with clinical features of persistent fetal vasculature (PFV), including cryptless iris, persistent pupillary membrane, centrally dragged ciliary processes, central posterior capsular opacity (accentuated around an enlarged Mittendorf dot), and iridohyaloid vessels. C10P had a unilateral total cataract in the right eye with PFV characterized by cryptless iris, persistent pupillary membrane, and bleeding vessels. C) Chromatogram of the LMW amplicon (∼370 bp) revealed direct splicing between exon 1 and exon 5, indicating loss of exons 2 to 4. D) In contrast, the control amplicon (∼697 bp) displayed normal splicing between exon 1 and exon 2. Red rectangular boxes beneath the chromatograms indicate the nucleotide sequences at the respective exon–intron junctions. The joined regions—exon 1 to 5 in the LMW transcript and exon 1 to 2 in the control transcript—are highlighted by blue boxes on the chromatograms.

To identify the possible splice site or other potential variant responsible for the loss of exons 2 to 4 in the CRYBA1 RNA transcript, we performed PCR followed by Sanger sequencing of all six exons of the *CRYBA1* gene using genomic DNA of the probands and their parents. None of the patients carried any coding, splice site, or splice region variants that could potentially impact normal splicing (Fig. 2A). Furthermore, to explore the possibility of deep intronic or large deletions that could have resulted in the absence of exons 2 to 4 in the transcript, we performed long-range PCR to amplify the full-length *CRYBA1* gene including exons 1 through 6 (∼7535 bp) (Fig. 2B) and sequenced it using Oxford Nanopore Technology. The binary alignment mapping (BAM) files were visualized using Integrative Genome Viewer. Interestingly, all the *CRYBA1* gene amplicons exhibited intact and complete genomic regions comprising exons 1 through 6, and none of the patients showed any deletions that could have led to the loss of exons 2 through 4 (Fig. 2C).

**Figure 2:**
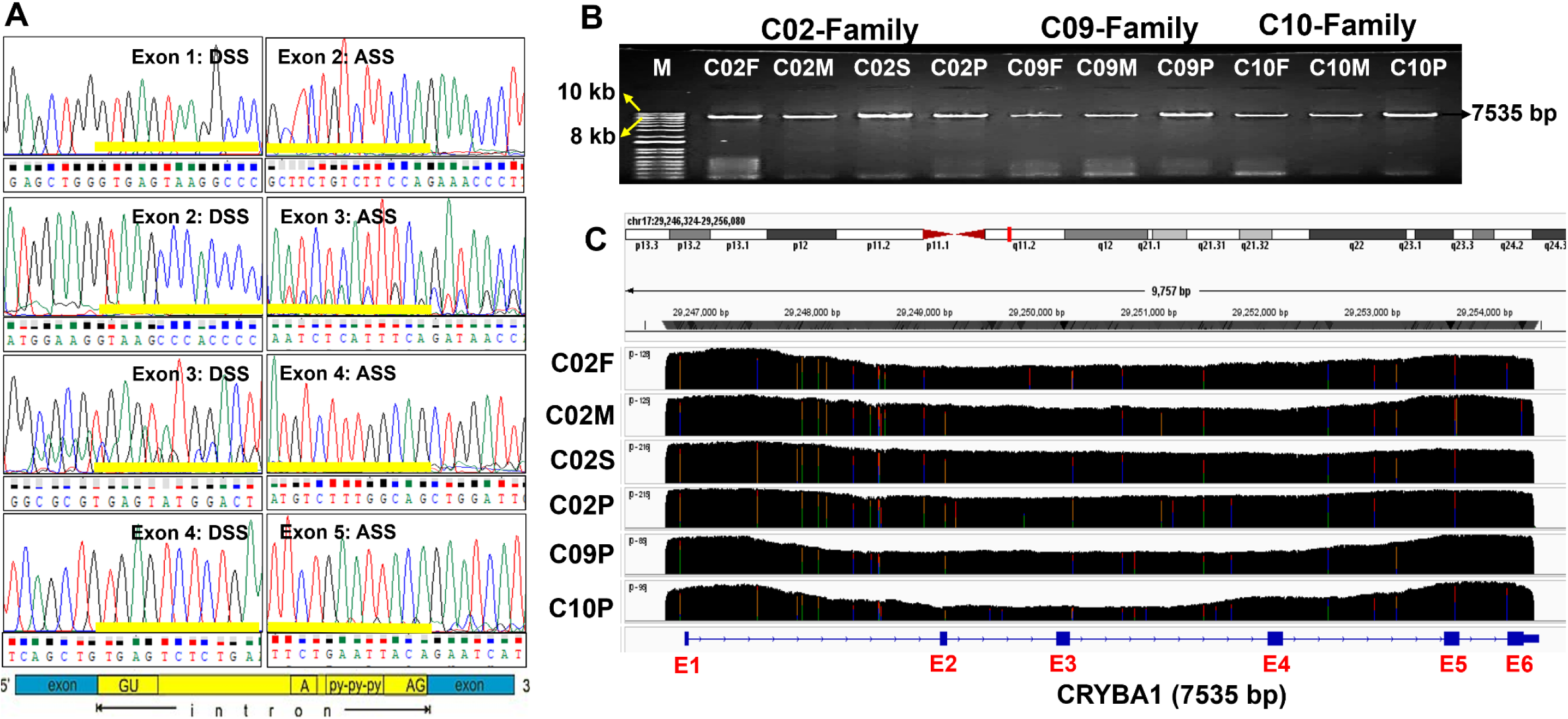
A) Chromatograms showing the splice regions of exons 1 through 5 of the CRYBA1 gene. Yellow bars highlight the donor and acceptor splice sites (DSS: Donor Splice Site; ASS: Acceptor Splice Site) at the respective exon–intron junctions. The bottom panel illustrates the canonical splice site sequences for reference. B) Agarose gel electrophoresis of the full-length CRYBA1 gene (from exon 1 through exon 6) displays the expected amplicon size of 7,535 bp from the probands (C02P, C09P and C10P) and their respective family members; M: 1kb DNA molecular weight marker. C) Integrative Genome Viewer (IGV) image of the CRYBA1 gene region encompassing exons 1 to 6. The top panel indicates the chromosomal location of the CRYBA1 gene. Mapped reads from the C02 family—father (C02F), mother (C02M), sister (C02S), and proband (C02P)—along with C09P and C10P, are shown as black extended bars. Coverage depth is noted to the right of each sample label, indicating the minimum and maximum read depth. Coloured vertical bars on the reads represent single nucleotide variants (green: adenine, blue: cytosine, saffron: guanine, red: thymine).

Upon long-range PCR and subsequent ONT sequencing, we found a total of 10 deep intronic variants in the *CRYBA1* gene common to all three probands C02, C09, and C10 (Table 4). Most of the variants were found in intron 1 (c.31+1013G>A, c.32-1085G>A, c.32-774C>T, c.32_550C>T, c.32-539G>A) and intron 3 regions (c.215+24C>T, c.216-568A>T, c.216-321C>T), as well as in intron 4 (c.357+400C>A) and exon 5 (c.456C>G) (Fig. 3A). The variant c.216-321C>T found in intron 3 was novel. The heterozygous variant c.32-1085G>A located in intron 1 is predicted to be mildly deleterious with a CADD phred score of 7.3 and is extremely rare (MAF<0.001) in gnomAD. Another heterozygous variant, c.357+400C>A located in intron 4, is predicted to cause alteration of splicing through activation of an intronic cryptic acceptor site. Furthermore, the heterozygous synonymous variant c.456C>G; p.Gly152Gly is predicted to create a new ESS site with a significant alteration in the ESE/ESS motifs ratio. We believe that the intronic variants c.32-1085G>A, c.357+400C>A, and c.456C>G, either individually or in various combinations, might influence the tissue-specific splicing process and the expression of AS-CRYBA1 transcript in the developing lens, particularly in these three probands.

**Figure 3:**
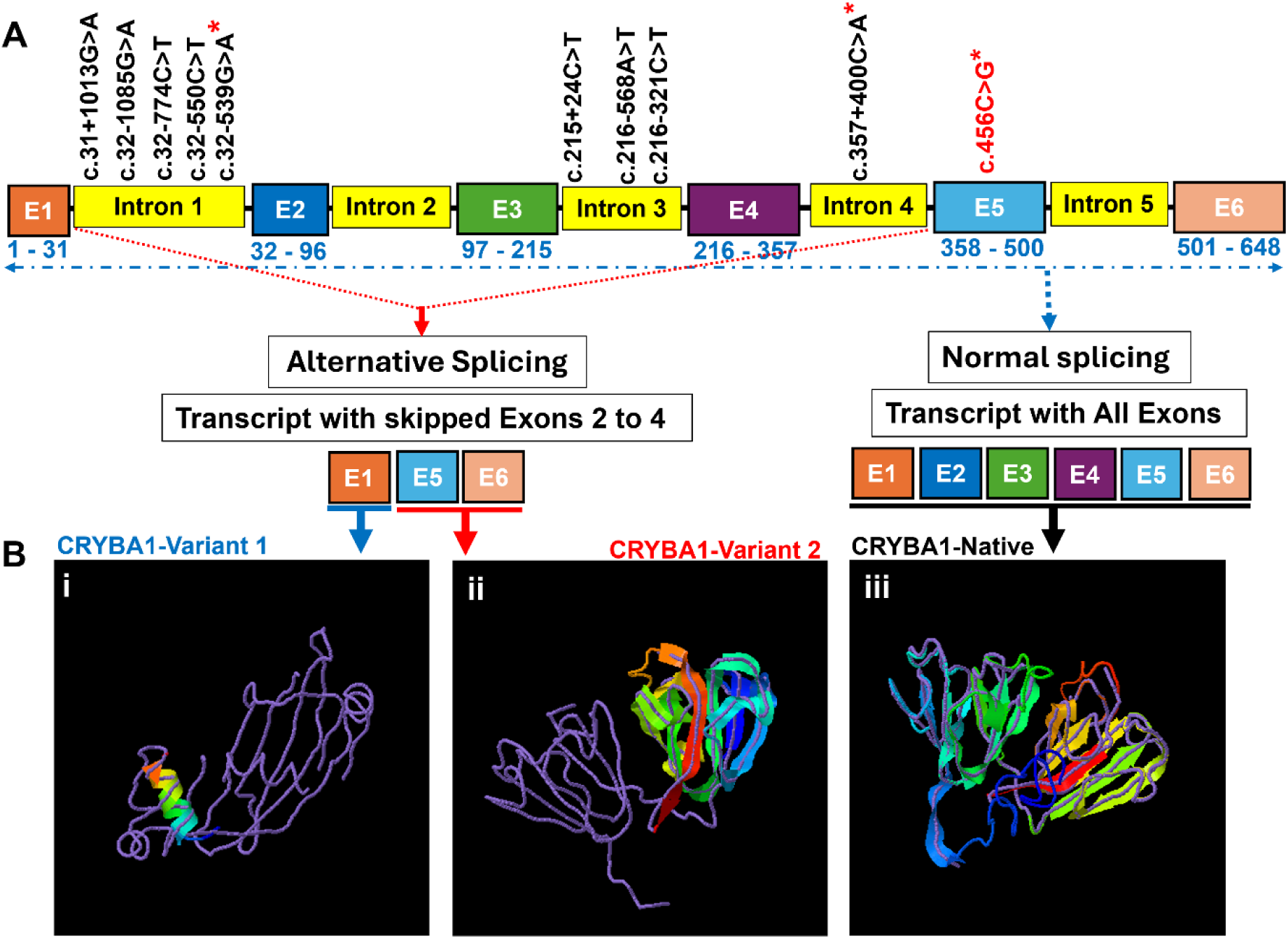
**A)** Cartoon illustrates the structure of the *CRYBA1* gene, including exons (E) and introns. Deep intronic variants common to all three probands are shown within each corresponding region, with the SNP marked by an asterisk (*) noted to influence splicing events. Coloured boxes represent exons (E1 to E6), and yellow-coloured boxes represent introns (1 to 5). The numbers below the exons indicate their length in base pairs. B) Illustration of alternative and normal splicing of *CRYBA1* transcript. Skipping of exons 2 to 4 is indicated by the red dotted line, resulting in two protein variants; i) CRYBA1-Variant 1 and ii) CRYBA1-Variant 2 of βA3/A1-crystallin. CRYBA1-variant 1 is translated from the native initiation site at Exon 1 (denoted by the blue line under the E1 box), while CRYBA1-Variant 2 is translated from a proximal initiation site located in exon 5 (denoted by the red line under the E5 and E6 boxes). iii) Normal splicing of the CRYBA1 transcript containing all six exons (indicated by blue dots and dashes), leading to the generation of the CRYBA1-Native protein of βA3/A1-crystallin (iii). In the images (i–iii), the purple strings represent the reference protein model (3lwkA), and the coloured strands correspond to the predicted protein variants. B(i) shows partial N-terminal domain of consists of first 11 amino acids translated from exon 1; B(ii) shows Greek key motifs 3 & 4 of last 90 amino acids translated from exons 5 and 6 using alternative translation initiation codon.

**Figure 4:**
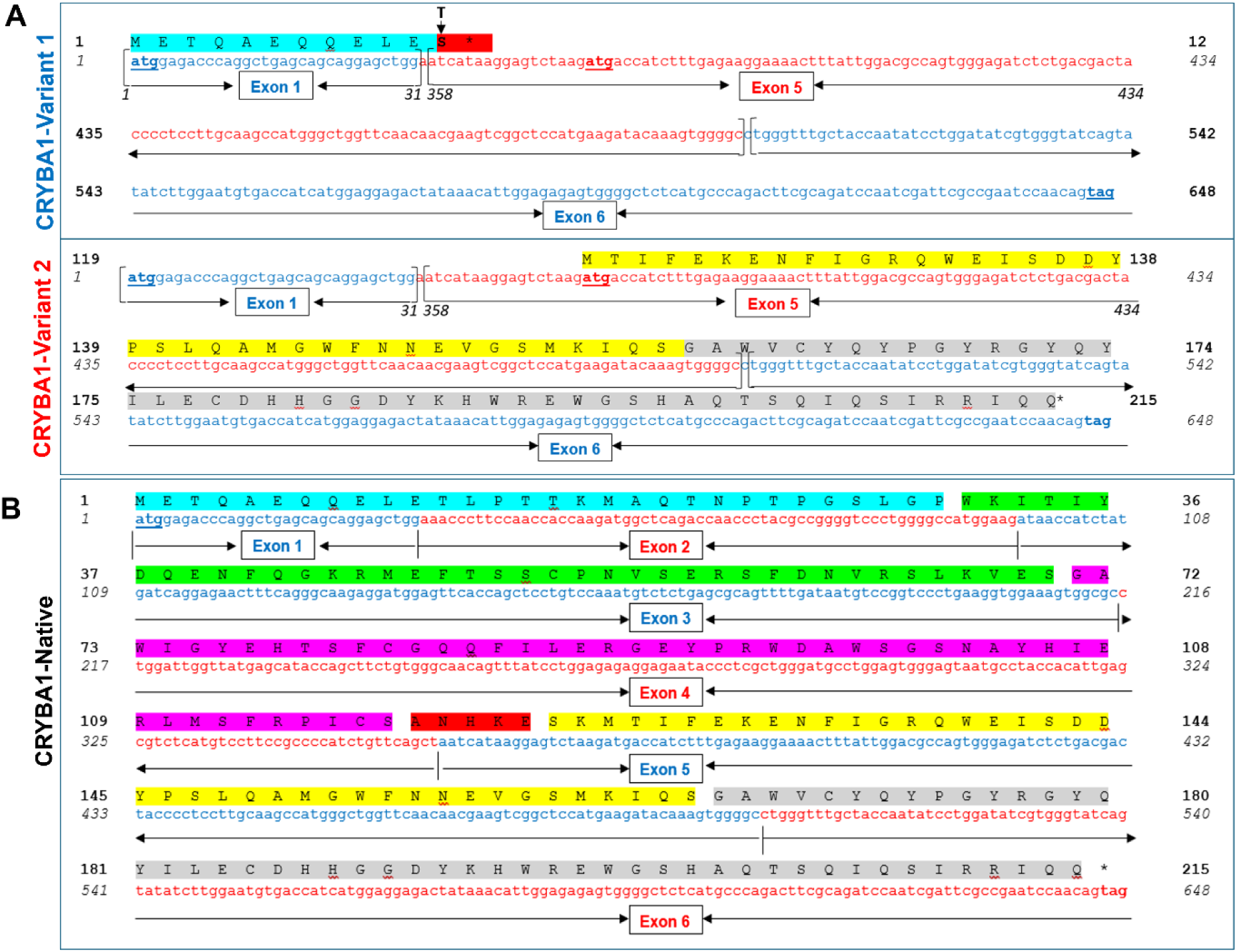
Illustration of nucleotide and corresponding amino acid sequence; **A)** CRYBA1 (variant 1), highlighted in cyan, is a truncated short peptide consisting of the first 11 amino acids of the N-terminal domain, resulting in a frameshift mutation (p.Thr12Serfs*2) and lacking all four Greek key motifs (1–4), assuming translation initiates from the first start codon. B) CRYBA1 (Variant 2), highlighted in yellow and grey, consists of 90 amino acids and lacks the N-terminal domain and the first two Greek key motifs (1 & 2), assuming translation begins from an alternative start codon located at nucleotide positions 50–52 (black arrowhead). C) CRYBA1 (Native) is a full-length βA3/A1-crystallin protein. Exons are displayed in alternating blue and red colours. Exon numbers are shown in boxes, and exon boundaries are indicated by arrows. Amino acid positions are marked in bold, while nucleotide positions are in italics. The colour shades represent various protein domains with corresponding nucleotide and amino acid positions: N-terminal domain (1–90; 1–30), Greek key motif 1 (91–210; 31–70), Greek key motif 2 (211–354; 71– 118), connecting peptide (355–369; 119–123), Greek key motif 3 (370–495; 124–165), and Greek key motif 4 (496–645; 166–215).

**Table 4:** List of deep intronic variants observed in CRYBA1 gene from ONT sequencing.

| Loci | Region | Variant | Zygosity | SNP ID | MAF | Human Splice Finder Analysis | CADD | ACMG Classification |
| --- | --- | --- | --- | --- | --- | --- | --- | --- |
| Chr17:29247907 | Intron 1 | c.31+1013G>A | Het | rs1389264911 | 0.000 | No significant splicing motif alteration | 5.2 | VUS (PM2, BP7) |
| Chr17:29248057 | Intron 1 | c.32-1085G>A | Het | rs1432755559 | 0.000 | No significant splicing motif alteration | **7.3** | VUS (BP7) |
| Chr17:29248368 | Intron 1 | c.32-774C>T | Het | rs120919158 | 0.000 | No significant splicing motif alteration | 0.1 | VUS (BP7) |
| Chr17:29248592 | Intron 1 | c.32-550C>T | Het | rs1033385934 | 0.000 | No significant splicing motif alteration | 1.3 | VUS (PM2, BP7) |
| Chr17:29248603 | Intron 1 | c.32-539G>A | Het | rs2459723 | 0.353 | <u>Alteration of an intronic ESS site</u> | 3.2 | B (BA1, BS2, BP7) |
| Chr17:29250324 | Intron 3 | c.215+24C>T | Het | rs780022667 | 0.000 | No significant splicing motif alteration | 0.5 | VUS (PM2, BP7) |
| Chr17:29251496 | Intron 3 | c.216-568A>T | Het | rs8075385 | 0.227 | No significant splicing motif alteration | 0.0 | B (BA1, BS2, BP7) |
| Chr17:29251743 | Intron 3 | c.216-321C>T | Het | Novel | N/A | No significant splicing motif alteration | 3.2 | VUS (PM2, BP7) |
| Chr17:29252605 | Intron 4 | c.357+400C>A | Het | rs8080081 | 0.291 | · <u>Activation of an intronic cryptic acceptor site (Potential alteration of splicing)</u> | **6.5** | B (BA1, BS2, BP7) |
|  |  |  |  |  |  | · Creation of an intronic ESE site |  |  |
| Chr17:29253738 | Exon 5 | c.456C>T:<br>p.G152G | Het | rs1047790 | 0.225 | <u>Creation of a new ESS site (Significant alteration of ESE / ESS motifs ratio (-10))</u> | 4.0 | B (BA1, BS2, BP6, BP7) |

Het: heterozygous; MAF: Minor Allele Frequency obtained from genomAD; ESE: Exonic Splice Enhancer; ESS: Exonic Splice Silencer; CADD refers to the Phred score from Combined Annotation Dependent Depletion (CADD), where a Phred score of 10 indicates that the variant is among the top 10% most deleterious variants, while a score of 20 places it in the top 1%, and so on.

Taken together, based on the observations from transcript and genomic sequencing, it is very likely that the observed loss of exons 2 through 4 in the CRYBA1 transcripts could be due to an alternative splicing (AS) mechanism. Given this possibility, we predicted two potential CRYBA1 protein variants based on the retained nucleotide sequences in the AS-CRYBA1 transcript (Fig. 3B & 4A). If the AS-CRYBA1 transcript undergoes translation using the native initiation site, it would produce a truncated peptide (p.Thr12Serfs*2; METQAEQQELE**S***\*) consisting of only the first 11 amino acids from the N-terminal domain and lacking all four Greek key motifs (CRYBA1-Variant 1 in Fig. 3B (i) & 4A). Alternatively, by using the second proximal translation initiation codon at nucleotide position 50–52 in the AS-CRYBA1 transcript, it would produce a small peptide consisting of 90 amino acids encoded partially by exon 5 and exon 6, lacking the N-terminal domain, the first two Greek key motifs, and the connecting peptide (CRYBA1-Variant 2 in Fig. 3B (ii) & 4B), and retaining Greek key motifs 3 and 4. 3D protein model of CRYBA1-Native revealed the presence of N-terminal domain, Greek key motifs 1 & 2, a connecting peptide, and Greek key motifs 3 & 4 (CRYBA1-Native in Fig.3B(iii) & 4B).

## Discussion

Detection of single nucleotide variations (SNVs) or indels from genomic DNA is a routine practice in genetic research and clinical diagnostics. However, RNA, being a highly dynamic molecule, undergoes context-specific changes such as alternative and aberrant splicing, fusion transcripts, allele-specific expression, and RNA editing [9]. Mutations in expressed genes, particularly driver mutations, are often conserved and more readily detectable at the RNA level [10]. Furthermore, mutation screening using RNA in large genes can be more efficient, requiring fewer PCR amplifications than DNA-based approaches [11]. Despite these advantages, RNA-based mutation detection remains underutilized in clinical settings due to challenges such as the requirement for tissue-specific samples [12–15].

In this study, we utilized lens epithelial and fiber cells from pediatric patients with unilateral cataracts to screen for transcript variants. We identified alternative CRYBA1 RNA transcripts producing low molecular weight amplicons (∼370 bp) in three probands. Sanger sequencing confirmed exon 1–5 junctions in these transcripts, indicating skipping of exons 2–4. Genomic DNA sequencing revealed no splice site variants. Long-range PCR followed by Oxford Nanopore sequencing showed intact CRYBA1 genomic regions but uncovered ten deep intronic variants common to all three probands. Among these, three variants (c.32-1085G>A, c.357+400C>A, and c.456C>G) are predicted to influence splicing regulation, suggesting the possibility of alternatively spliced CRYBA1 transcripts in human lenses.

The CRYBA1 gene comprises six exons spanning approximately 8 kilobases and encodes βA3/A1-crystallin. The first two exons encode a 32-amino acid N-terminal extension, while the remaining four exons encode four “Greek key” motifs. Sequence homologies among these exons and their intron-exon junctions support a model of gene evolution based on two successive exon duplications. Transcription of CRYBA1 in the eye lens initiates 24 base pairs downstream of a putative TATA box and just 7 nucleotides upstream of a potential start codon, resulting in a single mRNA transcript of approximately 1 kilobase [16]. This transcript can produce two polypeptides—βA3 and βA1-crystallin—using alternative translation initiation sites and a leaky ribosomal scanning mechanism that bypasses the first start codon and initiates translation at a downstream site. As a result, βA3-crystallin includes an additional 17 amino acids at the N-terminus compared to βA1 [17]. Structurally, βA3/A1-crystallin exhibits a conserved tertiary fold consisting of two homologous domains, each containing two symmetrical “Greek key” motifs formed by four-stranded antiparallel β-sheets. These domains are connected by a short linker peptide of 9–10 amino acids, contributing to the protein’s stability and functional integrity [18].

Based on in silico analysis, it was predicted that if the alternatively spliced CRYBA1 transcripts observed in this study undergo translation, they may generate two distinct aberrant βA3/A1-crystallin polypeptides (Figs. 3B & 4A). Firstly, as CRYBA1 variant 1 lacks part of N-terminal domain and all the Greek key motifs, it is likely that AS-CRYBA1 transcript may not be translated and instead might undergo nonsense-mediated decay (NMD) to prevent the production of truncated, potentially harmful proteins [19–22]. Secondly, as CRYBA1 variant 2, which lacks the N-terminal domain (that normally contains signal peptide), is unlikely to be secreted from the endoplasmic reticulum (ER), might accumulate in the ER and cause ER stress. Had the translated CRYBA1-Variant 2 secreted from the lumen of ER by other possible signal peptides, the loss of connecting peptide and Greek key motifs could disrupt βA3/A1-crystallin dimerization [23]. These mechanisms might inevitably impair lens protein turnover and compromise lens homeostasis via dominant-negative effects or cytotoxicity. It could be believed that despite the loss of first 2 Greek key motifs, the remaining polypeptide with two additional Greek key motifs (3 & 4) might play a role in mechanisms other than the development of lens structure. The possible reasons are one, the exons in CRYBA1 are evolutionarily duplicated genomic region, second, CRYBA1 is shown to express in tissues other than lens.

Mutations in the CRYBA1 gene are known to cause cataracts in humans and mice, highlighting the critical role of βA3/A1-crystallin in lens development and function [24–26]. Notably, most pathogenic mutations in CRYBA1 have been identified in the canonical donor splice site of exon 3 (e.g., c.215+1G>A) or involve small deletions (e.g., c.278–280delGGA, c.590–591delAG) [27–37], suggesting a vulnerability of CRYBA1 to disruptions in splicing and sequence integrity. While these mutations typically result in autosomal dominant cataracts with bilateral involvement, all three probands in this study exhibited unilateral cataracts present since birth. This discrepancy suggests that context-specific alterations in transcript structure, possibly influenced by deep intronic variants, may contribute to the asymmetry of disease manifestation. We thus believe that the manifestation of unilateral cataracts might be owing to the localized molecular mechanisms rather than the genetic contribution.

Interestingly, the probands also exhibited one or more associated anomalies such as persistent fetal vasculature (PFV), pre-existing posterior capsular defect (PPCD), and posterior capsular plaque (PCP), suggesting a potential link between AS-CRYBA1 transcript and these phenotypes. This association is supported by animal models. The Nuc1 rat, which carries a 27-bp insertion in exon 6 of CRYBA1, shows complex ocular defects including PFV [38,39]. Loss of functional βA3/A1-crystallin in these models results in astrocyte accumulation around the hyaloid artery and impaired lysosomal acidification in astrocytes, disrupting pathways such as mTOR and Notch/STAT3 [39,40]. Likewise, cryba3/a1 knockout mice display congenital cataracts and PFV due to defective lysosomal clearance and failure of hyaloid regression [41,42]. Ma and coworker also demonstrated that a c.215+1G>A splice site mutation in CRYBA1 led to the loss of exons 3 and 4 (c.97_357del), producing an unstable, toxic 15 kDa truncated protein (p.Ile33_Ala119del) that induced unfolded protein response (UPR), apoptosis, and lens capsule rupture in transgenic mice [43]. These findings corroborate our hypothesis that CRYBA1 transcript variants owing to the abnormal tissue specific molecular mechanisms may contribute to both cataract formation and associated anomalies such as PFV and PPCD.

In summary, we report the presence of alternatively spliced CRYBA1 transcripts in human lenses. Protein variants 1 and 2, which likely arise due to alternative splicing, may lack essential functional domains and structural integrity. While the precise molecular mechanisms underlying CRYBA1 dysfunction remain to be clarified, this study provides evidence for alternatively spliced CRYBA1 transcripts in pediatric patients with unilateral cataracts associated with PFV or posterior capsule defects. It also suggests a potential role for deep intronic variants in modulating transcript structure and function. To our knowledge, this is the first report documenting alternatively spliced CRYBA1 transcripts in human lenses. Future in vitro or in vivo studies expressing these transcript variants will be essential for understanding their pathogenic potential and functional relevance in the development of cataracts and associated anomalies such as PFV, PPCD, and posterior capsular plaque.

## Data Availability

All data produced in the present study are available upon reasonable request to the authors

## Acknowledgement

The authors extend their gratitude to the parents and siblings for their participation in this study.

## Author Contributions Statement

**Conception and design:** Sankaranarayanan Rajkumar, Abhay R Vasavada, Deepa Agarwal

**Analysis and interpretation of the data:** Sankaranarayanan Rajkumar, Deepa Agarwal, Shail A Vasavada

**Drafting of the paper:** Sankaranarayanan Rajkumar, Abhay R Vasavada

**Revising it critically for intellectual content:** Sankaranarayanan Rajkumar, Abhay R Vasavada, Vaishali A Vasavada

**Final approval of the version to be published:** All authors agree to be accountable for all aspects of the work.

## Data availability statement

The authors confirm that the data supporting the findings of this study are available within the article [and/or] its supplementary materials. Further queries can be addressed to the corresponding author.

